# Rapid systematic review on clinical evidence of chloroquine and hydroxychloroquine in COVID-19: critical assessment and recommendation for future clinical trials

**DOI:** 10.1101/2020.06.01.20118901

**Authors:** Yitong Wang, Shuyao Liang, Tingting Qiu, Ru Han, Monique Dabbous, Anna Nowotarska, Mondher Toumi

## Abstract

**Purpose:** This study aims to critically assess the published studies of Chloroquine (CQ) and hydroxychloroquine (HCQ) for the treatment of COVID-19 and provide recommendations for future clinical trials for the COVID-19 pandemic.

**Method:** A rapid systematic review was conducted by searching the PubMed, Embase, and China National Knowledge Infrastructure databases on April 13, 2020. Three clinical trial registry platforms, including ClinicalTrials.gov, the EU Clinical Trials Register, and the Chinese Clinical Trial Register were also complementarily searched.

**Results:** A total of 10 clinical studies were identified, including 3 randomized controlled trials (RCTs), 1 comparative nonrandomized trial, 5 single-arm trials, and 1 interim analysis. The heterogeneity among studies of the baseline disease severity and reported endpoints made a pooled analysis impossible. CQ and HCQ (with or without azithromycin) showed significant therapeutic benefit in terms of virologic clearance rate, improvement in symptoms and imaging findings, time to clinical recovery, and length of hospital stay in 1 RCT, 4 single-arm trials, and the interim analysis, whereas no treatment benefit of CQ or HCQ was observed in the remaining 4 studies. Limitations of the included studies ranged from small sample size, to insufficient information concerning baseline patient characteristics, to potential for selection bias without detailing the rationale for exclusion, and presence of confounding factors.

**Conclusion:** Based on the studies evaluated, there still lacked solid evidence supporting the efficacy and safety of HCQ and CQ as a treatment for COVID-19 with or without azithromycin. This emphasized the importance of robust RCTs investing HCQ/CQ to address the evidence uncertainties.

## Introduction

In late December 2019, a cluster of cases of pneumonia caused by 2019 novel coronavirus (2019-nCov) was first reported in Wuhan, China [1]. The virus was named Severe Acute Respiratory Syndrome Coronavirus 2 (SARS-CoV-2) by the International Committee on Taxonomy of Viruses (ICTV), and the disease was officially named Coronavirus Disease-2019 (COVID-19) by the World Health Organization (WHO). COVID-19 spread outside of China to such an extent that COVID-19 was declared a pandemic by WHO on March 12, 2020, and became a public health emergency of international concern [2]. As of April 13, 2020 (time of writing this paper), 1,872,073 confirmed cases were reported and 116,098 had died [3]. The COVID-19 pandemic presents an urgent challenge to identify effective drugs for treatment.

As of this writing, there is no proven effective pharmacologic treatment for COVID-19. Several agents are being tested in ongoing clinical trials [4], including chloroquine (CQ) and hydroxychloroquine (HCQ), lopinavir-ritonavir with and without interferon, remdesivir, tocilizumab, and convalescent plasma.

CQ, and its derivative HCQ, have recently gained attention because of their potential benefit in the treatment of COVID-19 [5]. As amine acidotropic forms of quinine for the treatment of malaria and chronic inflammatory diseases including systemic lupus erythematosus (SLE) and rheumatoid arthritis (RA), the sulphate and phosphate salts of CQ and HCQ have been used worldwide for more than 70 years and are part of the WHO model list of essential medicines. They are cheap and have an established clinical safety profile with generally mild and transitory side effects [6]. Recently, a large study covering more than 900 000 patients supported the safety of the product [7]. In vitro, CQ and HCQ were found to possess antiviral activity against RNA viruses as varied as rabies virus, poliovirus, HIV, hepatitis A virus, hepatitis C virus, influenza A and B viruses, influenza H5N1 virus, Chikungunya virus, Dengue virus, Zika virus, and Ebola virus, as well as DNA viruses such as hepatitis B virus and herpes simplex virus [8,9]. Recently, the in vitro antiviral activity of CQ and HCQ in inhibiting SARS-CoV-2 replication and their potential immunomodulating properties were reported [10,11]. One in vitro study suggested that HCQ might be more potent that CQ [11]. Mechanisms of action of CQ and HCQ may include inhibition of viral enzymes or processes such as viral DNA and RNA polymerase, viral protein glycosylation, virus assembly, new virus particle transport, and virus release.

CQ and HCQ are not approved by the US Food and Drug Administration (FDA) for the treatment of COVID-19. The FDA is, however, facilitating the availability of chloroquine during the COVID-19 pandemic to treat patients for whom a clinical trial is not available or participation is not feasible [12], and some protocols include the recommendation for the use of CQ and HCQ [13]. Several clinical guidelines published in China recommended the use of CQ (500 mg, twice per day, and for a duration no longer than 10 days) and HCQ (200 mg, three times per day) for the treatment of COVID-19 [14,13,15,16]. However, in France, HCQ was not recommended as an early treatment even though several clinical trials of HCQ were conducted by Raoult [17–19], a member of the expert council advising the president and government on the COVID-19 crisis.

Given the unclear efficacy and safety data in clinical studies and inconsistent recommendations in several countries about the use of CQ and HCQ for the treatment of COVID-19, the objectives of this study are to conduct a critical assessment of the published clinical trials of CQ and HCQ for the treatment of COVID-19 and provide recommendations for future clinical trials to fight the COVID-19 pandemic.

## Methodology

A rapid systematic literature review was conducted to identify clinical studies of CQ or HCQ for the treatment of COVID-19, following the guidance of rapid reviews to strengthen health policy and systems from World Health Organization (WHO).

A strategic search of 2 English language databases (Medline and Embase via the OVID interface), 1 Chinese language database (China National Knowledge Infrastructure [CNKI]), 3 clinical trial registry platforms (ClinicalTrials.gov, the EU Clinical Trials Register, and Chinese Clinical Trial Register) and Google Scholar for grey literature was performed using the keywords of “COVID-19, SARS-COV-2, new coronavirus, coronavirus disease 2019, chloroquine, and hydroxychloroquine” on April 13, 2020.

Clinical studies including randomized control trial (RCT), comparative non-randomized trial, single-arm trial, and interim analysis with published results in both English and Chinese were included in this study.

The following clinical study information was extracted: (1) study characteristics, including the authors, country in which clinical trial was conducted, starting and ending dates, eligibility and exclusion criteria, study design, primary outcomes, secondary outcomes, safety outcomes, and duration of follow-up for outcome assessment; (2) outcomes, including the dosage regimens for the intervention group and the control group, baseline characteristics of patients included in the intervention and control groups, and the results for primary outcomes, secondary outcomes, and safety outcomes.

The search of clinical trial registry platforms also garnered supplementary information regarding the study characteristics and outcomes of the included studies where needed. In addition, any inconsistencies between the methods reported in the publication and those in the protocol as described in the clinical trial database could be determined.

All processes of literature search, selection of clinical trials, data extraction, and critical assessment were conducted independently by two analysts. Any disagreements between two researchers were resolved by internal discussion and reconciled by a third, independent researcher.

A descriptive analysis of included clinical trials was conducted summarizing the study design and main results of published clinical trials investigating the effectiveness and safety of CQ or HCQ for the treatment of COVID-19.

Critical discussion of included clinical trials on the selection and measurement of outcomes and study quality in terms of evidence-based medicine assessment were conducted based on experts’ opinions to generate methodological recommendations for future clinical trials for the treatment of COVID-19, including objective, study design, population, endpoints, trial duration, statistical analysis and reporting.

## Results

A total of 10 published clinical studies of CQ/HCQ for the treatment of COVID-19 were identified, including 3 RCTs [20–22], 1 comparative nonrandomized trial [23], 5 single-arm trials [19,24,17,25,18], and 1 interim pooled analysis of clinical studies [26].

### Descriptive analysis of included clinical trials

The descriptive analyses including study characteristics and outcomes of included clinical trials are presented in Table 1a and Table 1b, respectively.

**Table 1a.**
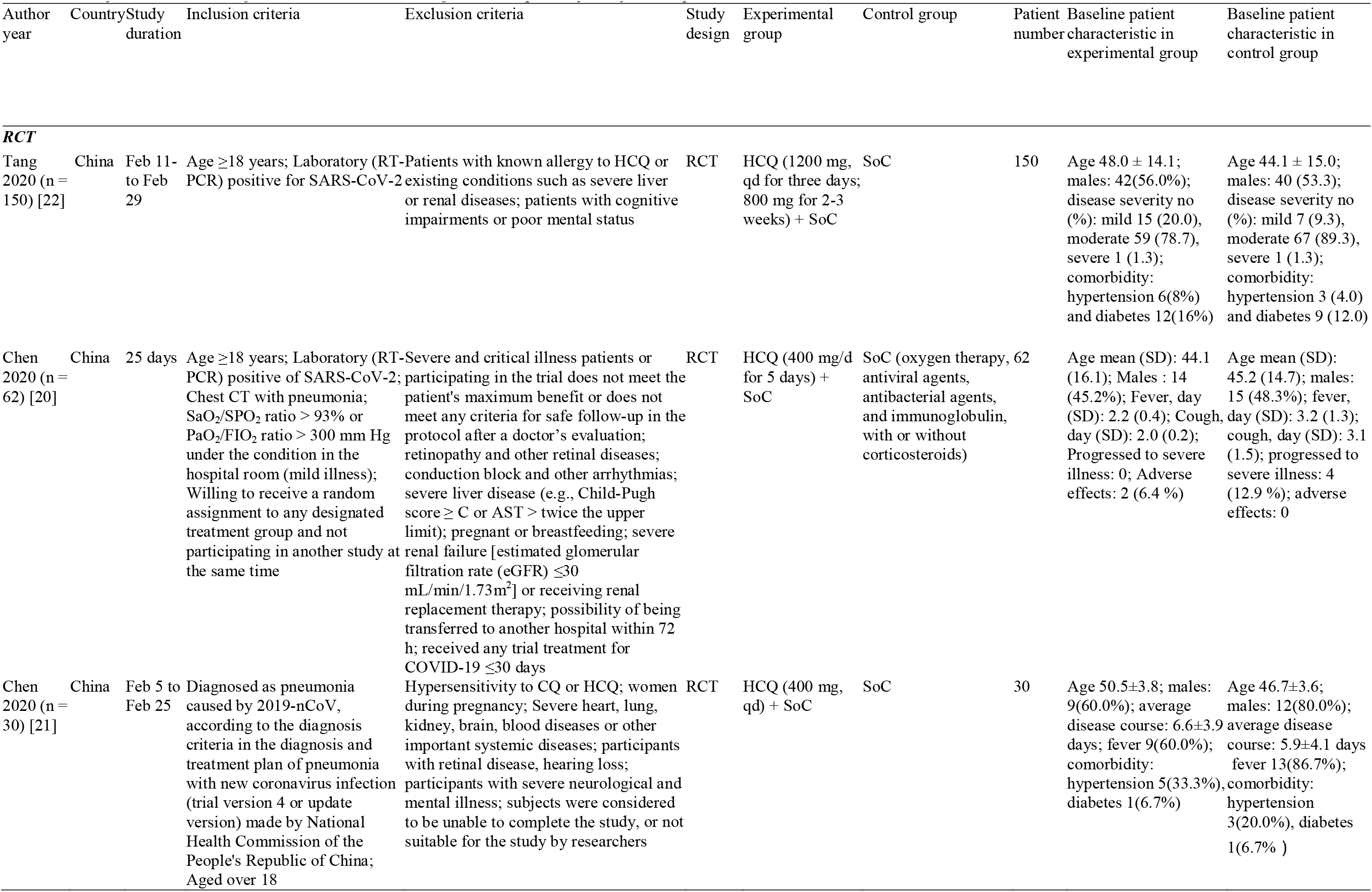
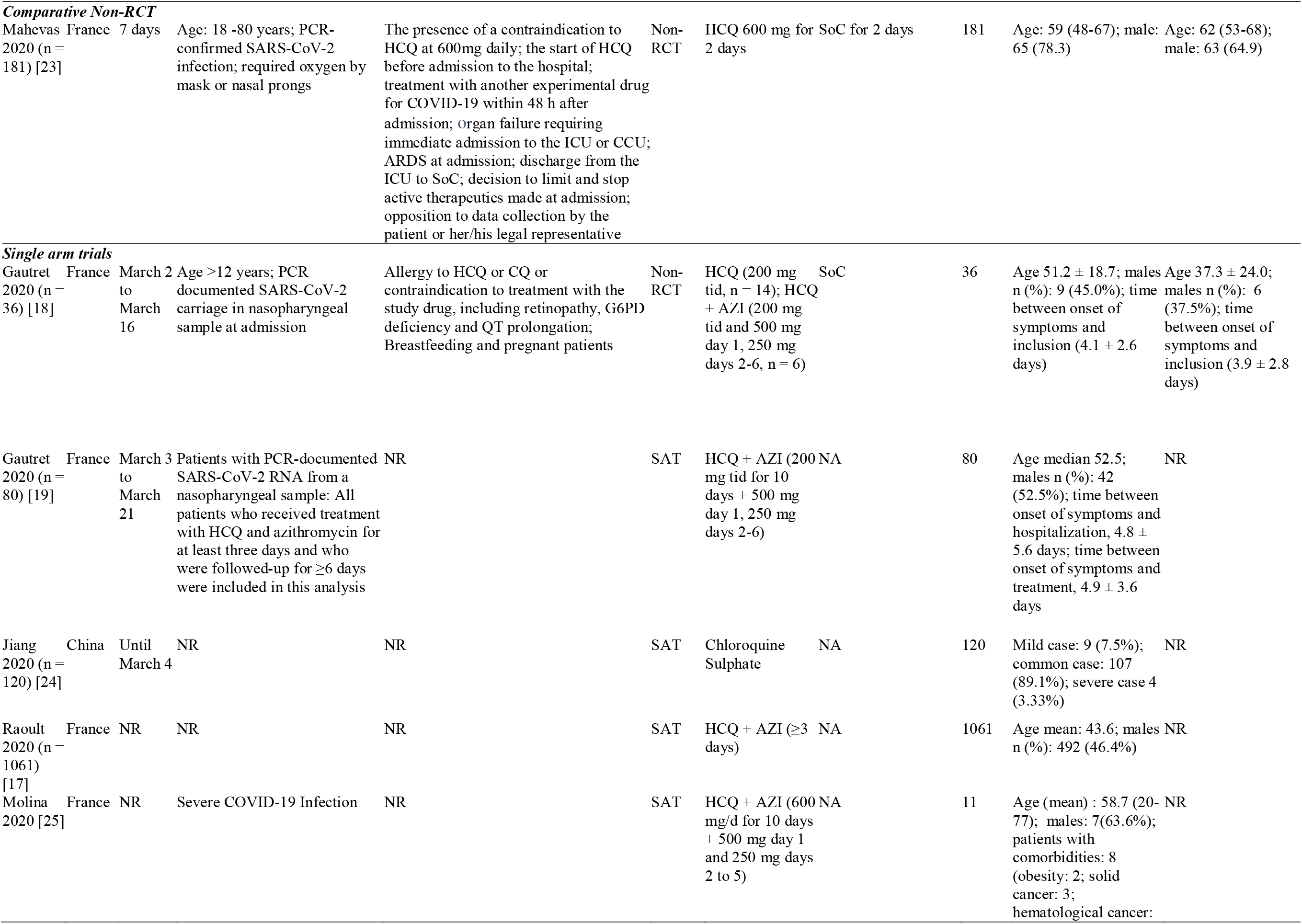
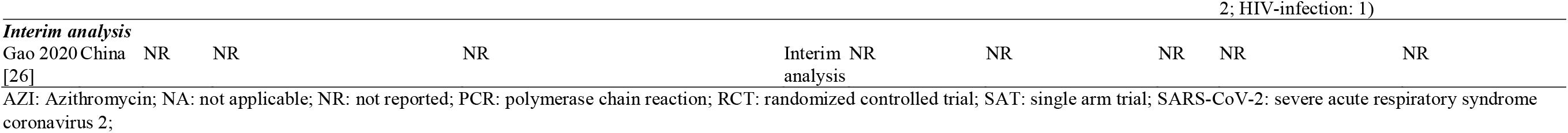
Study characteristics of included clinical studies for chloroquine/hydroxychloroquine

**Table 2b.**
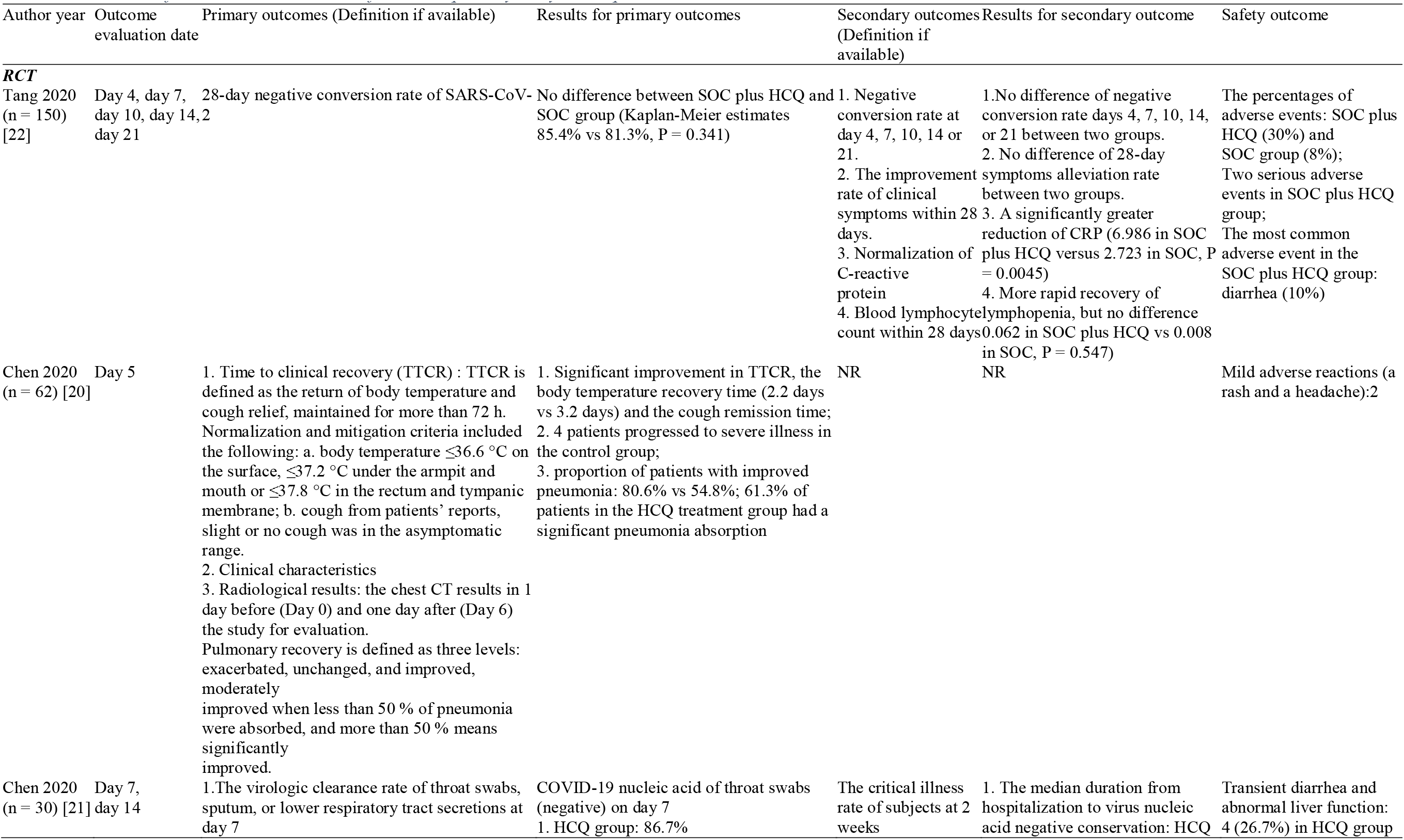
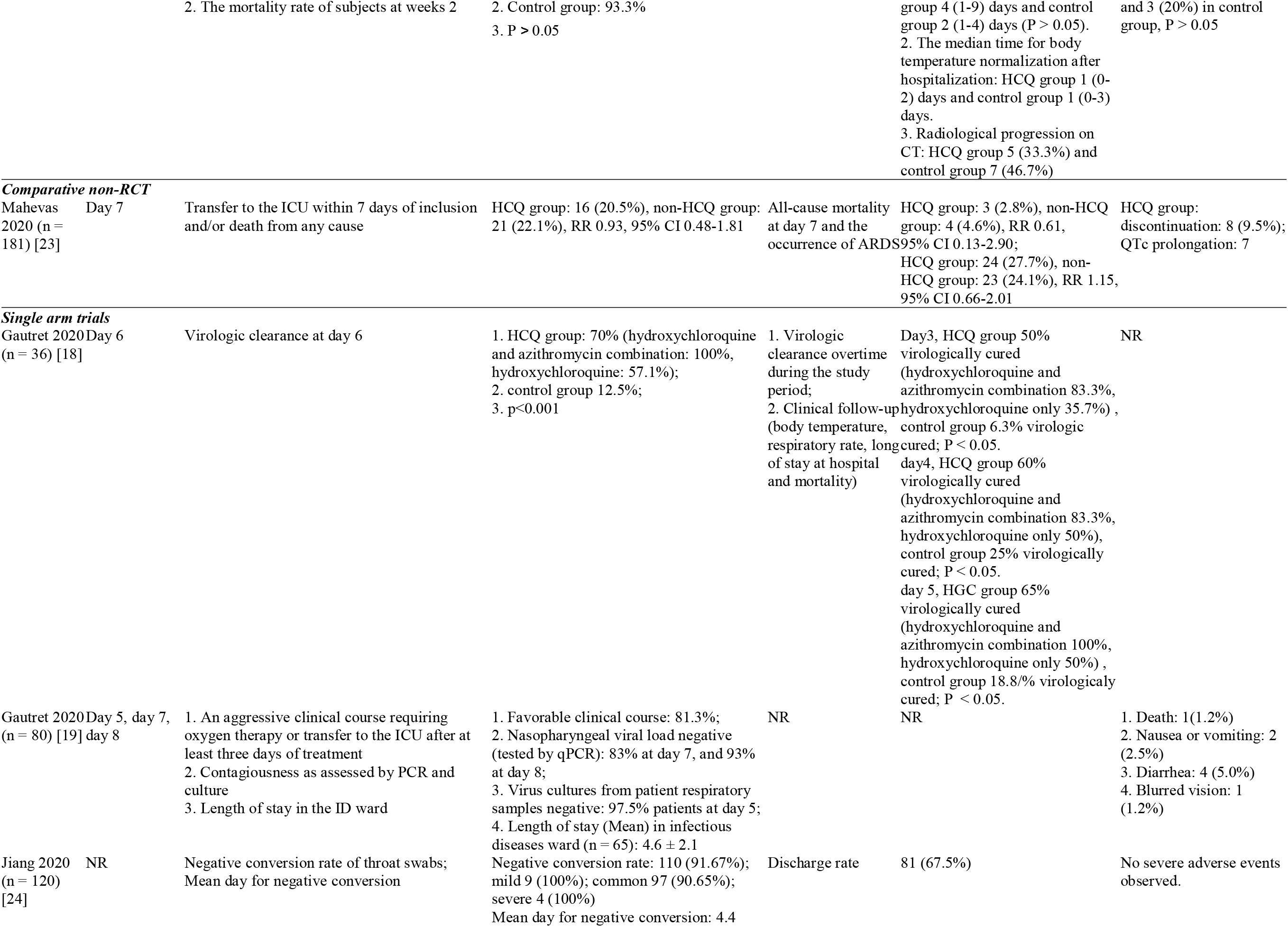
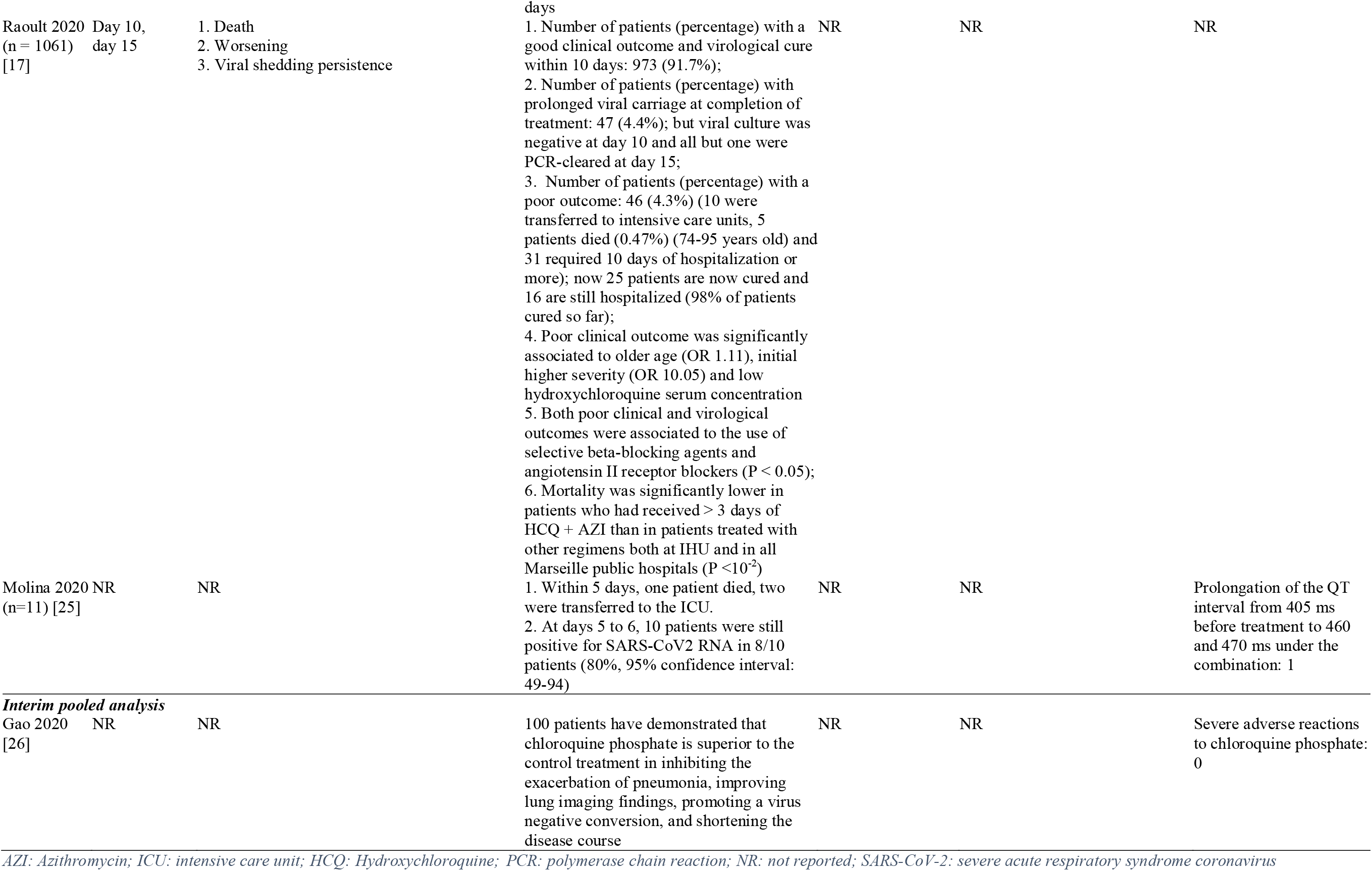
Outcomes of included clinical studies for chloroquine/hydroxychloroquine

#### Randomized clinical trials

##### Results from Tang et al (n = 150)

A randomized clinical trial of 150 patients assessed out of 191 screened patients (79.5%) was conducted by Tang et al [22]. The inclusion criteria were that patients be at least 18 years of age and have positive SARS-CoV-2 infection confirmed with real-time reverse-transcriptase–polymerase-chain-reaction (RTPCR). Patients were excluded if they were allergic to HCQ or had existing conditions such as severe liver disease, renal disease, cognitive impairments, or poor mental status. Patients were randomly assigned to the HCQ group (n = 75, HCQ 1200 mg/d for 3 days, then 800 mg for 2–3 weeks followed by standard of care [27]) and the control group using SoC (n = 75). The SoC was unrestricted, and all therapies were allowed. The primary end point (the 28-day negative conversion rate of SARS-CoV-2) and 4 secondary end points (negative conversion rate at days 4, 7, 10, 14, or 21, the improvement rate of clinical symptoms within 28 days, and normalization of C-reactive protein [CRP] and blood lymphocyte count within 28 days) were assessed and reported. For the HCQ group, the mean age (SD) was 48.0 (14.1) years and 42 (56.0%) were male. For the control group, the mean age (SD) was 44.1 (15.0) years and 40 (53.3%) were male. In the HCQ group, 6 of 75 did not receive HCQ: 3 patients withdrew consent and 3 patients refused to use HCQ. Patients in both groups received several antiviral therapies. No significant improvements in the 28-day negative conversion rate of SARS-CoV-2 were observed in HCQ group compared with the control group (85.4% vs 81.3%). No significant differences in negative conversion rates at days 4, 7, 10, 14, or 21 were observed in the HCQ group compared with the control group (the median negative conversion time: 8 days vs 7 days, P = 0.341). No significant differences in the rate of symptom alleviation within 28 days were observed in the HCQ group compared with the control group (59.9% vs 66.6%). A significant decrease of CRP value within 28 days was observed in the HCQ group compared with the control group (6.986 vs 2.723, P = 0.045). No significant differences in the blood lymphocyte count within 28 days were observed in the HCQ group compared with the control group (0.062 vs 0.008, P = 0.547). The post hoc analyses showed that no difference in the negative conversion time by the addition of HCQ upon SOC was observed in the analyzed subgroups (age, body mass index value, conditions, days between disease and randomization, baseline CRP value, contaminant use of potential antiviral agents and baseline lymphocyte count). However, the post hoc analysis after removing the confounding effects of antiviral agents indicated significant efficacy of HCQ in alleviating symptoms (hazard ratio, 8.83; 95% CI, 1.09 to 71.3). The rates of adverse events in the HCQ group and the control group were 30% and 8.8%, respectively.

##### Results from Chen et al (n = 62)

Another randomized clinical trial of 62 patients assessed out of 142 screened patients (43.6%) was conducted by Chen et al [20]. The inclusion criteria were a positive SARS-COV-2 diagnosis, pneumonia on computed tomography (CT) scan, and SaO_2_/SPO_2_ > 93%. Patients were excluded if their status was severe, or they presented with vital organ failure, or they had a contraindication to HCQ. Patients were randomly assigned to the HCQ group (n = 31, HCQ 400 mg/d for 5 days and SoC) and the control group (n = 31, SoC). Three end points (time to clinical recovery, clinical characteristics, and radiologic results) were assessed at baseline and day 5. For the 62 patients, the mean age was 44.7 years and 29 (46.8%) were male. No significant difference in age and sex distribution was observed between 2 groups. Significant improvements in time to clinical recovery were observed in the HCQ group compared with the control group (body temperature recovery time: mean [SD], 2.2 [0.4] vs 3.2 [1.3], P = 0.0008; cough remission time: mean [SD], 2.0 [0.2] vs 3.1 [1.5], P = 0.0016). A larger proportion of patients with improved pneumonia was observed in the HCQ group (80.6%) compared with the control group (54.8%). Of patients in the HCQ group, 61.3% had a significant pneumonia consolidation absorption. Two mild adverse reactions (1 report of rash and 1 of headache) were reported in the HCQ group.

##### Results from Chen et al (n = 30)

Chen et al [21] also performed a prospective, randomized study that enrolled 30 patients with COVID-19. Fifteen patients in the HCQ group were treated with the combination of HCQ (400 mg/day for 5 days) and SoC, including oxygen therapy, symptomatic treatment, and other antivirus drugs. The other 15 patients served as the control group and received SoC only. The primary outcomes included virologic clearance rate based on a respiratory pharyngeal swab 7 days after randomization and the mortality rate 2 weeks after randomization. The HCQ group and control group showed no significant difference in negative conversion rate of COVID-19 nucleic acid in respiratory pharyngeal swabs on day 7 (86.7% vs 93.3%), the median duration from hospitalization to virus nucleic acid negative conservation (4 days vs 2 days), or median time for body temperature normalization (1 day in both groups). A higher proportion of patients in the control group showed radiologic progression on CT images compared with HCQ group (46.7% vs 33.3%), but all patients showed improvement in the follow-up examination. All patients survived at the end of follow-up. No differences between the 2 groups in the incidence of adverse events was reported.

### Comparative nonrandomized trial

#### Results from Mahevas et al (n = 181)

Mahevas et al [23] collected the routine care data for COVID-19 patients in 4 French hospitals to simulate a targeted trial investigating the effectiveness and safety of HCQ 600 mg/day. Adult patients with PCR-confirmed COVID-19 who required oxygen were included, while patients who had contraindications to HCQ, received HCQ before admission, or received other experimental antivirus drugs were excluded from this study. An inverse probability of treatment weighting (IPTW) approach was used to emulate the randomization procedure and balance the baseline differences between treatment groups. The treatment outcomes for 2 strategies were compared: HCQ was given within 48 hours of admission (HCQ group) and no HCQ was given within the 48-hour period (non-HCQ group). The primary outcome was a composite of admission to the ICU within 7 days after inclusion and/or death due to any causes. All-cause mortality at day 7 and the incidence of acute respiratory distress syndrome (ARDS) were studied as secondary outcomes. A total of 181 patients were investigated, 84 in the HCQ group and 97 in the non-HCQ group. Patient baseline characteristics were well balanced between the 2 groups, except for a higher rate of confusion in the non-HCQ group. Seventeen (20%) of the patients in the HCQ group received concomitant azithromycin. A marginally lower percentage of patients in the HCQ group were transferred to ICU or died compared with the non-HCQ group (20.2% vs 22.1%), but no significant differences were shown between the groups. Three (2.8%) patients died within 7 days of admission in the HCQ group compared with 4 (4.6%) patients in the non-HCQ group, and 27.4% of patients experienced ARDS in the HCQ group compared with 24.1% of patients in the non-HCQ group. Eight (9.5%) patients in the HCQ group discontinued HCQ due to the ECG modifications after a median of 4 days of treatment.

### Single arm trials

#### Results from Gautret et al (n = 36)

A prospective, open-label, nonrandomized controlled trial was conducted by Gautret et al [18] with the inclusion of 36 confirmed COVID-19 patients. Patients were included if they were over 12 years old and had PCR-documented SARS-CoV-2 carriage in nasopharyngeal samples at admission regardless of their clinical status. The exclusion criteria included allergy to HCQ or CQ or other known contraindications to treatment such as retinopathy, G6PD deficiency, and QT prolongation, or were breastfeeding or pregnant. The intervention arm was 26 patients who received oral HCQ 200 mg, three times per day for 10 days; the remaining patients did not receive HCQ and served as negative controls. Among the active treatment patients, 6 patients also received azithromycin (500 mg on day 1, followed by 250 mg per day for the next 4 days) for the prevention of bacterial superinfection. The primary end point was the clearance of SARSCoV-2 carriage in nasopharyngeal specimens after 6 days of inclusion. At day 6 of inclusion, the negative conversion rate of COVID-19 nucleic acid for the active treatment patients was 70% compared with 12.5% in the control group (P = 0.001). Among the active treatment patients, 100% of the patients treated with the combination of HCQ and azithromycin tested SARS-CoV-2 RNA–negative in nasopharyngeal swabs compared with 57.1% in patients treated with HCQ alone (P < 0.001).

#### Results from Gautret et al (n = 80)

Based on this preliminary clinical trial of 36 patients [18], a further observational study with a larger sample size (n = 80) and longer follow-up duration (at least 6 days) was conducted by Gautret et al [19]. Patients received the combination of HCQ (600 mg/day for 10 days) and azithromycin (500 mg on day 1 and 250 mg on days 2–6) for the treatment of COVID-19. Three main end points (clinical outcome, contagiousness as assessed by PCR and culture, and length of stay in the infectious diseases [ID] unit) were reported in the study. A total of 80 patients (median age, 52.5 years; male/female, 42/38) were included; 79 of the patients received treatment on a daily basis for no more than 10 days and 1 patient stopped treatment on day 4 for the potential risk of intervention with another drug. Sixty-five (81.3%) patients had a favorable clinical outcome and were discharged from the unit. Three patients were transferred to the ICU, of whom 2 improved and returned to the ID ward. One patient died in the ID unit. 15% of patients used oxygen therapy during the treatment period. 83% and 93% of patients tested negative for nasopharyngeal viral load by qPCR at day 7 and day 8, respectively. 97.5% of patients tested negative on virus cultures from respiratory samples at day 5. Of the 65 patients who were discharged from the ID ward, the mean (SD) time from initiation to discharge was 4.1 (2.2) days and mean (SD) length of stay was 4.6 (2.1) days.

#### Results from Jiang et al (n = 120)

Jiang et al [24] led a single-arm study in the respiratory department of Sun Yat Sen Memorial Hospital. A total of 120 patients with COVID-19 were included and treated with chloroquine phosphate until March 4, 2020. The dosage and treatment duration of chloroquine phosphate were not reported in the publication. In all, 89.1% of patients were moderate cases, 7.5% showed mild symptoms, and 3.33% had severe status. At the end of study, 110 patients tested negative for viral nucleic acid in throat swabs after the treatment. Among the negative cases, all mild and severe cases were virologically cured, and 90.65% of moderate patients were cured. The average time to negative conversion was 4.4 days. None of the 120 patients receiving chloroquine phosphate developed critical COVID-19, and 81 patients had been discharged. During the study period, no serious adverse reactions were found.

#### Results from Raoult et al (n = 1061)

A cohort study of 1061 COVID-19 patients assessed out of 3165 positive patients was conducted at IHU Mediterranean Infection by Raoult et al [17]. Patients who were treated with the combination HCQ and azithromycin (HCQ-AZ) for at least 3 days and followed up for at least 9 days were included. Three end points (deaths, worsening of condition, and persistence of viral shedding) were reported. For the 1061 patients included, the mean age was 43.6 years and 492 (46.4%) were male. A total of 973 (91.7%) patients had a favorable clinical outcome and achieved virologic cure within 10 days, and 46 patients (4.3%) had a poor clinical outcome, of whom, 10 patients were transferred to ICU, 5 died, and 31 needed more than 10 days of hospitalization. 98% patients were cured so far. Forty-seven patients (4.4%) had prolonged viral carriage at the end of treatment, which was associated with the high viral load at diagnosis (P < 0.01). However, all 47 of these patients tested negative for viral culture at day 10, and 46 tested clear by PCR at day 15. Poor clinical outcome was significantly associated with older age (odds ratio [OR], 1.11), greater severity (OR, 10.05), low hydroxychloroquine serum concentration, and use of selective beta-blocking agents and angiotensin II receptor blockers. Patients who received more than 3 days of HCQ-AZ treatment had a lower mortality rate compared with patients treated with other drugs in all Marseille public hospitals (P < 0.01).

#### Results from Molina et al (n = 11)

An article by Molina et al [25] gave a short report on the outcome of 11 consecutive COVID-19 patients admitted in the department of infectious diseases and treated with HCQ (600 mg per day for 10 days) and azithromycin (500 mg on day1, and 250 mg for the next 4 days). The criteria for inclusion and exclusion of patients were not reported in the study. The average age for the patients was 58.7 years. The baseline characteristics of included patients showed 8 patients with significant comorbidities that may have affected their prognosis. Of all the patients, 1 required oxygen therapy, 2 were transferred to the intensive care unit (ICU), 1 patient died, and 1 patient discontinued after the appearance of QTc interval prolongation, which was considered a serious adverse event during the treatment. Of the patients in this study, 80% did not reach nasopharyngeal viral clearance at the end of study.

### Interim pooled Analysis

#### Results from Gao et al (n = 100)

Gao et al [26] reported the narrative results of an interim analysis of COVID-19 patients who received treatment with chloroquine phosphate from different clinical trials. The authors reported that for more than 100 patients, chloroquine phosphate was associated with a significant improvement in reducing viral load, controlling pneumonia as assessed by CT scan, and shortening of disease course. The authors also reported 15 clinical trials conducted in China to investigate the efficacy and safety of CQ or HCQ in the treatment of COVID-19 pneumonia. Of the 15 trials, 8 were currently recruiting as planned, 2 trials were now pending, and 5 trials were cancelled by the investigators. Of the 10 noncancelled clinical trials (Table 3), 4 were to investigate the treatment effects of chloroquine phosphate, 5 were to investigate the treatment effects of HCQ, and 1 was to investigate the preventive effect of HCQ on close contacts after exposure to COVID-19; most trials (n = 7) were RCTs with sample sizes ranging from 78 to 320 patients; time to RT-PCR negativity (n = 5) was the most common primary end point.

**Table 3.** Clinical trials of Chloroquine Phosphate and Hydroxychloroquine Sulphate

| Trial ID | Planned date of enrollment | Recruitment status | Study design | Population (number of patients) | Intervention (dosing, if available) | Control (dosing, if available) | Primary outcome |
| --- | --- | --- | --- | --- | --- | --- | --- |
| ChiCTR2000029939 | 10/02/2020 | Recruiting | RCT | COVID-19 pneumonia (n = 100) | Chloroquine phosphate + standard of care | Standard of care | Days of hospitalization |
| ChiCTR2000029935 | 06/02/2020 | Recruiting | Non-RCT | COVID-19 pneumonia (n = 100) | Chloroquine phosphate+ standard of care | NA | Days of hospitalization |
| ChiCTR2000029609 | 10/02/2020 | Pending | Non-RCT | COVID-19 pneumonia (n = 146) | Mild-moderate patients:<br>Group 1: chloroquine phosphate<br>Group 2: lopinavir-ritonavir<br>Group 3: chloroquine phosphate + Lopinavir-ritonavir<br>Severe patients:<br>Group 1: chloroquine phosphate<br>Group 2: lopinavir-ritonavir | NA | Time to nucleic acid negativity |
| ChiCTR2000029899 | 17/02/2020 | Recruiting | RCT | COVID-19 pneumonia (n = 100) | Hydroxychloroquine sulfate (day 1: first dose: 600 mg, second dose: 600 mg after 6h; day 2~5: 200mg, bid) | Chloroquine phosphate (day 1-3: 500 mg, bid; day 4-5: 250 mg, bid) | Time to clinical recovery |
| ChiCTR2000029542 | 03/02/2020 | Recruiting | Non-RCT | COVID-19 pneumonia (n = 20) | Chloroquine phosphate | Standard of care | Time to nucleic acid negativity; Mortality |
| ChiCTR2000029559 | 31/01/2020 | Recruiting | RCT | COVID-19 pneumonia (n = 300) | Group 1: hydroxychloroquine (0.1 g, bid)<br>Group 2: hydroxychloroquine (0.2 g, bid) | Placebo | Time to nucleic acid negativity |
| ChiCTR2000029740 | 11/02/2020 | Recruiting | RCT | COVID-19 pneumonia (n = 78) | Hydroxychloroquine (0.2 g bid) | Standard of care | Time to nucleic acid negativity |
| ChiCTR2000029868 | 10/02/2020 | Recruiting | RCT | COVID-19 pneumonia (n = 200) | Hydroxychloroquine sulfate | Standard of care | Time to nucleic acid negativity |
| ChiCTR2000029898 | 17/02/2020 | Recruiting | RCT | Severe COVID-19 pneumonia (n = 100) | Hydroxychloroquine sulfate (day 1: first dose: 600 mg, second dose: 600 mg after 6 h; day 2-5: 200 mg, bid) | Chloroquine phosphate (day 1-3: 500 mg, bid; day 4-5: 250 mg, bid) | Time to clinical improvement |
| ChiCTR2000029803 | 14/02/2020 | Pending | RCT | Close contacts after exposure to COVID-19 pneumonia (n = 320) | Group 1: Hydroxychloroquine, small dose<br>Group 2: Hydroxychloroquine, high dose | Group 1: abidol hydrochloride, small dose<br>Group 2: abidol hydrochloride, high dose | Number of patients who have progressed to suspected or confirmed within 24 days of exposure to new coronavirus |

*The standard of care followed the guidance of the "Diagnosis and Treatment Protocol for COVID-19 Pneumonia" published by the Chinese National Health Commission[13].*
*Nucleic acid negativity is defined by reverse transcription polymerase chain reaction (RT-PCR)*
*Time to clinical recovery is defined as the normal body temperature (underarm temperature $\leq 36.9$ degrees C) and cough relief (evaluated by the cough symptom score scale) in the duration of at least 72 hours from the start of study treatment*
*RCT: Randomized clinical trial; COVID-19: Coronavirus Disease 2019; NA: Not available; bid: twice daily*

## Discussion

### Critical Discussion of the Published Studies

#### Randomized clinical trials

In vitro evidence has generated interest in HCQ and CQ as treatments for COVID-19. However, for virologic infection it is well established that in vitro studies may not be predictive of clinical benefit [10,11]. It can just be the starting point of hypothesis testing but not relevant for hypothesis generation.

However, it is not worth discussing HCQ/CQ in vitro experiments when a substantial number of humans in vivo clinical trials are available. So far, results from 3 randomized trials are available [21,20,22], as well as 1 nonrandomized comparative trial and 5 observational studies [24,17–19,25,23]. In addition, an interim analysis of 15 heterogeneous trials, not all randomized [26], pooled on the first 100 available patients have been reported as a narrative, with no further report on a larger sample.

##### Discussion for Tang et al (n = 150)

The study from Tang et al [22] was negative, but in this study patients were eligible for any possible concomitant therapies, leading to wide use of antiviral therapies in both arms. When adjusted for antiviral treatment, however, the rate of clinical benefit was higher in the HCQ arm but not statistically significant. These results are very difficult to interpret because the details of the concomitant antiviral therapies are not described and the distribution in both arms is not reported. Moreover, patients appear to be more severely ill in that study than in in Chen and al. [19]

##### Discussion for Chen et al (n = 62)

Of the 3 RCTs [21,20,22], one [20] on 62 patients is positive, showing a clear benefit of HCQ for all clinical end points and pneumonia as assessed by CT scan. Unfortunately, results are short term (assessment at day 6) and not reported beyond). Moreover, the study did not give a clear disposition of nonincluded patients, who constituted 56.4% of patients. At the least, it is important to understand how many were excluded because of a contraindication to chloroquine. This would have shed light on the proportion of COVID-19 patients for whom chloroquine is not appropriate. The 2 treatment arms were reasonably comparable even though the HCQ patients tended to have severe status. More patients had fever and were coughing in the HCQ group compared with the SoC group. End points were days of fever and cough and evolution of pneumonia as assessed by CT scan at the end of the treatment phase (day 6) compared with day 0. Patients on HCQ had fewer days of fever and cough, which was statistically significant but not substantial, but the rate of pneumonia improvement was 80.7% for the HCQ group compared with 54.8% for the SoC group. Pneumonia in 6.5% of patients was exacerbated in the HCQ treatment arm compared with 29% of patients in the SoC arm. The study’s limitation is the short-term duration of patient follow-up as it stopped at day 6. At the time of study end, 19.4% of HCQ-treated patients showed no improvement or exacerbation of pneumonia, and 14.9% were moderately improved. It would be very useful to have a clear view of longer term improvement and vital status of the patients. This was the first published study providing randomized evidence of the benefit of HCQ in COVID-19 patients as compared with SoC.

##### Discussion for Chen et al (n = 30)

The third randomized trial [21] on 30 patients tested HCQ on top of SoC. The protocol allowed all antiviral therapies to be used as SoC. Moreover, patients on CQ may have received interferon. Therefore, it does not seem to have been appropriately designed to test the benefit of HCQ/CQ. This significantly limits the interpretation of the results and makes it difficult to use for decision making. Ultimately all randomized controlled trials were compared with SoC. SoC happened to allow any therapy including several antiviral therapies.

#### Comparative nonrandomized trial

##### Discussion for Mahevas et al (n = 181)

Mahevas et al [23] reported a nonrandomized comparative study with HCQ augmented with azithromycin in a sample of patients. It is unclear how these patients were selected to receive azithromycin. Although differences are not statistically significant, a numeric trend in favor of HCQ did appear. Three (2.8%) patients died in the HCQ group compared with 4 (4.6%) patients in non-HCQ group within 7 days of admission. The study population was quite severely ill given they were hospitalized when hospitals were overloaded and only admitting the most severe cases with established pneumonia or severe condition. This figure deserves interest nonetheless.

#### Single-arm trials

In addition to these randomized trials, there have been 5 single-arm trials [18,19,24,25,17] published. One study [17] is only available as an abstract, but it included detailed patient dispositions at baseline and study end. One study (11 patients) [23] had negative results while the other 4 studies (with 1061, 120, 80, and 36 patients) showed great treatment benefit from HCQ or the combination of HCQ + azithromycin [17,24,19,18]. These studies may carry some limitations especially that the potential for selection bias is high. However, examination of the results supports that the intervention is likely effective. The lack of comparative arms also makes interpretation difficult.

##### Discussion for Gautret et al (n = 36)

Gautret et al [18] reported a single-arm study of poor quality that assessed the benefit of HCQ with and without azithromycin. In a post hoc analysis the authors performed a comparison versus a reference population but it cannot be used to support any decision analysis because of several selection biases. The limitations of the study have been widely commented on [28,29]. This study is the only one to document the additional benefit of adding azithromycin to HCQ compared with HCQ alone. The combination resulted in an increase in viral clearance from 55% to 100% at day 6, depending of subgroup analysis. The very small size and the several biases make it difficult to translate its findings to the broader population, but it is a signal that should not be ignored.

##### Discussion for Gautret et al (n = 80)

Gautret et al [19] reported a study of the combination of HCQ and azithromycin in 80 patients, but it is similar to a later study published by Raoult et al [17] on 1061 patients even though patients did not overlap. Therefore, we will only comment on the similar but much larger study as it provides more information on a much larger sample size.

##### Discussion for Jiang et al (n = 120)

The single-arm study led by Jiang et al [24] showed rapid viral clearance as early as day 4.4 on average. The population had mostly mild illness, but the improvement rate was not stratified by severity even though the number of severely ill patients was very small. This study supports the potential benefit of CQ; however, as with any noncomparative study, selection biases could not be excluded.

##### Discussion for Raoult et al (n = 1061)

Raoult et al [17] reported that out of 3165 patients who tested positive for SARS-CoV-2 less than a third (1061 patients) were included in the study. Because the inclusion criteria were very broad, it is important to understand the causes of noneligibility for the excluded patients, as well as their outcomes. This critical information will help determine whether there was no recruitment bias and whether patients at higher risk had not been excluded from the selected cohort. At first glance, the population identified [17] seems representative of the French population based on various criteria such as average age (41.8 vs 43.6 years [30]); male sex (48% vs 46.4% [31]); and prevalence of cancer (1.2% to 1.8% vs 2.6% [32]), diabetes (5% to 6% vs 7.4%[33]), coronary artery disease (4% vs 4.3% [34]), hypertension (30.6% vs 14% [35]), COPD (7.5% vs 10.5% [36]), and obesity (6.5% vs 5.8% [35]). 95% of patients had a low National Early Warning Score, a scale used to identify acutely ill patients [37]. It was medium in 2.4% and severe in 2.6% of patients. This suggests an overall low severity of illness in the studied population. 34.3% of patients had a normal pulmonary CT scan, and 22.7% had clearly established pneumonia (medium to severe imaging results), consistent with the finding that 2.6% of patients were severely ill according to National Early Warning Score classification. Hydroxychloroquine was at infratherapeutic plasma concentration (≤ 0.1 µg/mL) in 11.4% of patients. Of these patients, 13.3% had a good outcome and 40% had a poor outcome. Although concomitant medications were associated with poorest outcome, it may also be a confounding factor related to concomitant disease. Thus, the potential impact of treatment should be adjusted based on comorbidity to identify which comorbidity or treatment was the risk factor and, eventually, if both happen to be independent risk factors, what the relative weight is of each factor. Nasal viral load was used to confirm SARS-CoV-19 carriage, but no sensitivity and specificity are available for the technique used. 91.7% of patients were cleared of viral load at day 10, and almost all patients were cleared at day 15. This suggests the time to viral load clearance should not be more than 15 days. The rate of intensive care unit admission (1%) was low, and the mortality rate was also low (0.5%). The age range for the dead patients was from 75 to 95 years. Out of the 46 patients with poor outcomes, 5 deaths occurred, there were 10 hospitalizations in the ICU, and there were 31 hospitalizations of 10 days or more. The mortality rate among patients with poor outcomes was 11%, and the cure rate was 53% [17]. Overall, the results look very favorable, compared with current death rates of 6.3% [38] globally, the lowest being in Germany and Korea at about 2% [39]. That comparison may be difficult, however, because the denominator depends on the intensity of the testing. Nonetheless, in France, the mortality rate excluding those occurring in nursing homes is 9.1% and for hospitalized patients it is 13.2%, and the proportion of cured patients is 39% in hospitalized patients, as compared with 53% in this study [40]. The testing campaign at Marseille IHU was larger than in France as a whole, which is supported by the rate of positive testing results in France compared with the Marseille IHU cohort (29% and 8.2%, respectively). The rate of testing is 3.5 times higher in Marseille than in France in general. If we adjust the death rate based on the highest testing activity at Marseille IHU, the death rate would be 1.65%. This is still below those of Korea and Germany and much below the general French death rate. The data support use of HCQ, and it is expected on the final publication that, if selection bias can be excluded, these data will be very convincing in support of the value of hydroxychloroquine combined with azithromycin in treating COVID-19 patients. However, the role of each of drug in the combination remains quite uncertain. Only one study, with a very small sample size, compared azithromycin combined with HCQ versus HCQ alone, but there was channeling based on indication that is clearly reported.

##### Discussion for Molina et al (n = 11)

Molina [25] reported the outcome of 11 consecutive COVID-19 patients admitted to the infectious diseases department and treated with HCQ (600 mg/d for 10 days) and azithromycin (500 mg day 1 and 250 mg days 2 to 5). Average age was 58.7 years. At baseline 8 patients had significant comorbidities that might already have affected the patient’s prognosis. All but one required oxygen therapy, 2 were transferred to the ICU, 1 patient died, and 1 patient discontinued because of a serious adverse event (QTc interval prolongation). 80% of patients did not achieve viral clearance based on nasopharyngeal samples at study end. None of the patients had a plasma concentration of HCQ below the efficacity threshold.

This series is rather small and is inconsistent with the other observational studies. The actual severity of the patients with COVID-19 symptoms is insufficiently described, but they may well match the population of Chen et al [21] [20]—hospitalized COVID-10 patients with pneumonia. The severity of patients’ comorbidities might also be a differentiating factor between the studies with positive and negative results.

#### Interim Pooled Analysis

##### Discussion for Gao et al (n = 100)

Gao et al [26] reported the narrative results of 15 clinical trials conducted in 10 hospitals in Wuhan to assess the benefit–risk of HCQ and CQ in COVID-19 patients with pneumonia. The authors reported on more than 100 patients, gathered from the 15 studies. A reduction in viral load, control of pneumonia as assessed on CT scan, and shortening of disease course were observed among patients administered active treatment (HCQ or CQ) compared with patients who were administered SoC. These findings led to a “consensus” conference on February 15, 2020, followed by a new recommendation introducing HCQ and CQ in all subsequent guidelines as the treatment of choice for COVID-19 in China. This report was extensively discussed in China, but the information published is very scarce. This study should be considered a signal to the scientific community that HCQ and CQ work and should be considered as effective treatment options. It is very unfortunate that the further results for the 15 clinical trials were not reported in detail, as was certainly the case for the conference of February 2020. Although it is a strong signal, it could not be considered as evidence for an evidence-based medicine assessment. Ignoring it, however, would be reckless. Table 3 shows the heterogeneity of the studies pooled.

#### Methodological recommendations for future trials for COVID-19

##### Objective

The objective concerning the intervention under evaluation should be clearly established. It may aim to (1) reduce disease transmission; (2) prevent disease progression and fatalities in patients with mild-to-moderate infection, including at-risk patients; or (3) treat severely ill patients, including patients in the ICU. The objective will clearly affect the design, the end points to be used, the length of the study, and obviously the patients to be enrolled in the clinical trial.

##### Study design

A comparative trial is always preferred, and randomization ensures comparability of the assessed treatment arms. If researchers opt for a single-arm study, they should track the characteristics of patients not included at time of screening and their outcomes for the key study end points. In addition, in the case of a single-arm study, a comparison with a matched historical or, if feasible, prospective cohort receiving SoC would be desirable. A clustered randomization design would be better than a matching-adjusted comparison. This would entail patients being treated with SoC giving informed consent and all centers agreeing not to administer active treatment to patients if they are randomly assigned as controls, which is very unlikely within the current context of the COVID-19 pandemic.

A very high rate of patient refusal to enter a trial could be expected for a randomized blinded trial using a placebo intervention. Patients knowing the high risk may be willing to try anything even if not approved and of unknown benefit but not be willing to risk receiving the placebo. Thus, a blinded study may not be successful. A comparative trial to SoC might not work either as patients randomized to SoC will likely ask to drop from the study and require an active treatment.

In comparative studies, when mortality is the primary end point (an event such as mortality or discharge), an event-driven (number of events) trial design would be preferred. Indeed, in that case, the study enrollment would end when several events occurred and not when a given number of patients had been recruited to the study. We need to define a minimum expected difference between an active and a reference arm and from that assess the number of events required reach a statistically significant difference and power the study accordingly.

Because of the great uncertainty surrounding this new disease and the potential benefit of interventions that will be tested, the standard study designs based on deterministic statistics appear to be inappropriate. Indeed, the sample size calculations in these trials require information on the magnitude of the differential benefit (even if driven by assumptions) and the distribution of the end point estimate. Although interim analyses are possible, they would not be appropriate as they would consume power to adjust for α inflation and would delay time for results. Given the uncertainty involving sample size calculations, the risk of inconclusive outcomes remains too high to be acceptable in the light of the emergency situation. It might also be unethical to perform such a study because of the risk of negative study results. If researchers decide to maintain the deterministic statistics for their study, they should consider a 2:1 or, better still, a 3:1 (active: control) randomization design. From an ethical perspective it is very difficult to see how an ethical committee could allow such a comparative trial designed based on deterministic statistics and how they would develop their rationale.

On the other hand, the adaptive design [41] appears to be well suited. It allows adjustment over time for the included population profile and the sample size without inflating type I error. It would be conclusive under all circumstances, whether positive or negative. It allows assessment on an ongoing basis, at predefined intervals, of the probability of success of the intervention or, eventually, of the need to stop the study because of low chances of success. It is, in terms of quality, the type of design that all decision makers would require in this situation. It would prevent an inconclusive trial, expose only the required number of patients, adjust toward the appropriate population, and provide signals and direction on an ongoing basis.

##### Population

For assessing reduction of disease transmission, asymptomatic and mildly symptomatic patients are the ideal population. To assess impact on disease prognosis, asymptomatic or mild to moderate patients at risk of complications could be included, as well as all patients with very early and early stages of pneumonia. Patients with more severe illness and those at advanced stages may not respond to the same therapy because of the massive cytokine release described with COVID-19. Those severely ill patients may require other therapies because antivirals might not reach the inflamed tissue. Obviously, the population will depend on both the objective of the study and the type of drug tested.

a. To test for reduction of time to transmission, an asymptomatic population in whom the virus is identified by PCR could be used. In those cases, drugs able to eliminate the virus such as antiviral therapies would be the obvious intervention. Supportive therapy to stimulate patient immune defense might also be appropriate in that population.
b. To assess the impact on disease prognosis, targeting patients at risk or patients with early stages of pneumonia would allow a reduction in the number of patients needed for the trial and in the trial duration. In such a case antiviral treatment would be the target, with or without an agent that may mediate the immune response to target the potential cytokine storm.
c. To treat patients with pneumonia, a trial should target patients with moderate stages of pneumonia rather than severe cases. It is expected that if a therapy is effective with moderate severities of pneumonia, all patients would thereafter receive it, reducing the incidence of severe cases. In that case, antiviral therapies might not be effective because of the potential problems with cytokine storm, bacterial overinfection, and hypoxemia. Thus, treatment should include an immunomodulator able to control the cytokine storm. Antibiotics would likely be part of SoC; however, comparison of different antibiotic classes might be considered on top of immunomodulators. Therapy likely to enhance oxygen transport might also be considered, if available.
d. Patients in ICU might require all usual vital function interventions. At that stage, immunomodulatory treatment may be tested in clinical trials but the most important goal would be maintenance of vital functions. Obviously, comparing different immunomodulators might be an option, and single-arm comparisons would be acceptable, but placebo-controlled trials might not be an option.

Clearly in stages c and d it is very difficult to consider a placebo-controlled or SoC comparative trial. It might not be acceptable from an ethical perspective. In the ICU when there is sufficient clinical experience to suggest the benefit of an intervention, it should be used, particularly when the vital prognosis of the patient is involved.

For the situations outlined in a and b, a placebo-controlled trial might be an option; however, if they progress, patients should be able to withdraw from the trial, their treatment should be unblinded, and existing effective treatment should be administered based on the benefit–risk evaluation. Each patient is unique and should be treated that way.

##### End points

To reduce disease transmission, the time to clearance of viral load is an appropriate end point. For assessing risk of progression, clinical assessment (fever, cough, etc) and imaging (chest CT scan) appear to be the appropriate end points. Hospital admission and ICU admission are often biased end points because they are very dependent on the local infrastructure, the local clinical practice, and the importance of the number of patients that may dramatically overload the health care facilities. It may be an appropriate end point in a single-center study, but the practice may evolve over time and become an inappropriate end point for single-arm studies. Cure and mortality will also be important end points. For more severely ill patients, mortality and cure are the most important end points. Time to discharge is also an important end point, but hospital overcrowding may play a role in time to discharge. This may not be a major bias in randomized trials.

In all cases, antibody assessment should be continued for a long period after cure and at least until the viral carriage is negative.

Opting for 2 coprimary end points may also be a solution for some studies as it will ensure a stronger outcome and each end point may overcome the limitations of the other one.

##### Trial duration

Although treatment duration is short, patient follow-up should be long enough, with at least 1 visit 1 month after end of treatment, to assess the clearance of all symptoms and any potential adverse events. It is advisable whenever possible to follow up on antibody development and sustained response over a longer period of time, potentially 1 year. Because outcomes of the study are unknown 1 year of follow-up on potential coagulation, cardiac, and mainly respiratory function is a strong recommendation. Some authors have suggested a risk of pulmonary fibrosis even among the mildest cases. This risk is too severe to be ignored in the follow-up of clinical trials. It may be disease related or treatment related, including, for example, a high concentration of inspired oxygen (FiO_2_). However, high FiO_2_ may not explain the occurrence of pulmonary fibrosis if its presence is confirmed in patients with mildest symptoms not receiving oxygen.

##### Statistical analysis

For an adaptative design study, use of Bayesian analysis is recommended for the primary or coprimary end point(s). However, for secondary end points deterministic statistics can be used. Since there are usually several secondary end points, the issue of multiplicity of tests becomes a concern. Testing each end point separately at an *α* ≤ 0.05 will inflate the type I error. Methodologists would likely recommend having a fixed-sequence method [42], also called hierarchical statistical analysis, for the secondary end points. This in fact is a totally theoretical point in the current circumstances and should not be considered unavoidable.

For single-arm studies with an external comparative arm, propensity score matching is appropriate and should be used to minimize the biases. Alternative methodologies for matching-adjusted comparisons are possible and should be described and justified. The most challenging issue will be defining the matching criteria, and a deep analysis of risk factors for the considered primary end point will be required. Using a design of 1 patient on active treatment to 3 or 5 control patients would allow a reduction in the number of patients in the single arm and the duration of such a trial.

Subgroup analyses should be considered as well as risk factors for positive or negative outcomes. If these have not been prespecified in the statistical analysis plan, it may be considered hypothesis testing but may prove useful to support decision making, given the state of emergency of this situation. In such analyses, confounding factors may be numerous and should be accounted for whenever possible.

A committee blinded to the results and data might be established to define the confounding factors for which adjustment would be necessary.

##### Reporting

Extensive reporting should be made available as supplementary material of publications to ensure the scientific community transparent access to all information in order to allow for a scrutiny of the evidence generated and the opportunity to fully share the insights from the published studies.

#### What do we know beyond these trials?

Researchers from Wuhan University published a report on the use of HCQ and found that among 80 patients with systemic lupus erythematosus who were treated with hydroxychloroquine, none of them were infected with SARS-CoV-2 during the outbreak. HCQ might hold promise of protective effects for patients against COVID-19 [43].

In China, CQ was first recommended as one of the mainstream treatments for COVID-19 in the National Diagnosis and Treatment Protocol for Novel Coronavirus Pneumonia (6th trial version) on February 19, 2020 [44]. It could be an important indicator that the treatment potential of CQ was acknowledged at the national level. Thereafter, several influential regional guidelines released by different provinces also included CQ or HCQ for treatment of COVID-19 [45]. The Shanghai experts’ consensus for integrative treatment of COVID-19 [15] highly recommended HCQ as first choice before CQ and other antiviral drugs. Researchers in Shanghai have concluded that HCQ can effectively and safely prevent progression of disease based on the fact that since February 5 the incidence of new severe and critical cases of COVID-19 in Shanghai has decreased significantly after treatment with HCQ [46]. In the provinces in which regional guidelines are not established, CQ, one treatment recommended in the national guideline, will be considered the standard treatment. Analysis of the experience of clinical treatment in sharply decreasing the infection rate and fatality rate from China, sorting hospitals based on utility, differentiating patients and their treatments based on patients’ conditions, and integrating traditional Chinese medicine into therapy demonstrates that these approaches are practical and successful [47]. Clinical practice and scientific research were combined to strengthen the selection of effective drugs. Such drugs as chloroquine phosphate, favipiravir, and plasma from recovered patients went into clinical trials and produced timely feedback.

Among other Asian countries, guidelines drafted by the COVID-19 Central Clinical Task Force in South Korea in February 2020 recommended CQ or HCQ for the treatment of COVID-19 in elderly patients or patients with comorbidities [48]. The India Council of Medical Research (ICMR) recommended the prophylactic use of HCQ for asymptomatic health care workers or household contacts of suspected or confirmed COVID-19 cases [49].

Doctors in some countries have been given the option to use CQ and HCQ in hospitals to treat COVID-19 patients, which indicates efficacy of HCQ and CQ. On March 28, 2020, the FDA issued an Emergency Use Authorization (EUA) of hydroxychloroquine sulfate and chloroquine phosphate products supplied from the Strategic National Stockpile (SNS) for certain hospitalized patients with COVID-19. This announcement was not a genuine authorization but allowed doctors to prescribe it to adolescent and adult patients hospitalized with COVID-19, as appropriate, when a clinical trial is not available or feasible [50]. On April 1, 2020, the European Medicines Agency has announced that CQ and HCQ can be used for the treatment of COVID-19 in connection with clinical trials or in national emergency use programs for the treatment of COVID-19 patients [51]. Accordingly, European countries, such as France, Germany, and Denmark, allowed the use of CQ and HCQ on an experimental basis. In Italy, the L. Spallanzani National Institute for the Infectious Disease published their recommendations for HCQ or CQ combined with another antiviral agent for the treatment of COVID-19 [52]. The Polish National Drug Agency has amended the CQ specification and authorized it for supportive treatment of coronavirus such as SARSCoV-2 [53].

Some recommendations or guidelines around the world, including African countries such as Morocco and Tunisia [54], have proposed HCQ or CQ as treatment options with more or fewer precautions and left it to physicians to decide according to their appreciation of the benefit–risk.

#### Why the recommendation was not taken in several countries

The debate about chloroquine has become personal and has polarized the scientific community for and against HCQ, neglecting to analyze the facts within their context and ignoring the Chinese randomized clinical trial. The negative randomized study has been used as an argument against HCQ on the sole basis of the abstract, with limited effort to analyze the full content.

It is the state of the art to request double-blind randomized trials to assess the benefit risk of an intervention. In emergencies such as this, blinding is not an option as it would require several months to produce the study drugs according to GMP with a sufficient stability.

Randomization remains an obvious requirement, and all clinical trials should be randomized to ensure comparability of the treatment arm and thereby lend a solid internal validity of the study outcome. However, the FDA and European Medicines Agency have under several circumstances recognized a positive benefit–risk ratio based on a single-arm trial. This is obviously a suboptimal situation, but it did not prevent regulators from making decisions.

In that very polarized and political situation, the debate has turned dogmatic, between 2 opposing visions of medicine and scientific research. When methodologists require gold standard evidence as the only path to recommend an intervention, they ignore evidence-based medicine which recommends use of all available evidence and grades the strength of the evidence used to support the decision. Case series from clinician experience can be used to make policy recommendations in that methodology.

Serendipity has led to a number of major discoveries, including Pasteur and vaccination, Fleming and penicillin, Curie and radioactivity. Although proper methodology guarantees robust evidence generation, it should not be used as dogma when critical decisions must be made. This situation brings us back to the parachute trial vs placebo describe by Smith and Pell [55], two great methodologists who warned against the dictates of a dogmatic methodology that represents the only and unique truth. Their conclusion about the randomized blinded parachute trial is very clear. Only 2 options exist: (1) under exceptional circumstances, assess the potential risks and benefits of interventions, or (2) common sense might be applied. We continue our quest for exclusive evidence-based interventions and preclude any intervention used outside constraints of double-blind randomized trials. By the same criteria, we could say that Columbus did not discover America as it was not the hypothesis he tested when leaving Castilla in 1492, in the Santa Maria, to find an alternative route to India. It was just a hypothesis generation trip, while the second trip in 1493 was a conclusive hypothesis testing experience. Therefore, America was discovered by Columbus in 1493 not 1492. We would then have to rewrite the history. It seems that we all forget the lesson and focus on 2 dogmas, on one side that of the methodologists, and on the other side that of the infallible clinician’s experiences. Common sense may not be that common in times of crisis among either experts or policy decision makers, making this situation worrisome. Where is the pilot?

#### Conclusion

The lesson from all these trials is very interesting. In China, all randomized studies versus SoC have been confounded as SoC included several antiviral therapies. This prevented several studies from showing a differentiation from SoC. There is a question to raise about this bias: is it a methodological mistake? Certainly not. The guidelines have recommended several antiviral therapies and, early on, immunomodulators. Therefore, the physicians used their usual SoC. The study would thus examine whether a given intervention performs better than the usual SoC, whatever it may be. It would not confirm if one antiviral therapy is better than no antiviral therapies. This was not the question being asked by the Chinese experts, and doctors would not do anything less than give their patients the best option as a SoC in such an emergency situation for the patients, for the health care system, and for the society as a whole. Altogether, however, these studies suggest that at early stages of the disease, patients with mild illness might benefit from CQ or HCQ. The lack of granularity in the data reported prevents us from reaching firm conclusion.

Observational studies showed the value of combined HCQ and azithromycin. Per evidence-based medicine criteria, use of this combination would be recommended, but it would be weak. Therefore, we are still in urgent need of a RCT in mild to moderate severity COVID-19 patients to conclude on the benefit risk of HCQ and CQ with our without supplementation with azithromycin.

The vehement debate over the pros and cons of CQ and HCQ has demonstrated how emotional and dogmatic the scientific community may become in a crisis. The disagreement that has emerged among experts about valid treatments and their inability to achieve common ground about a recommendation with the necessary sense of urgency casts doubt on all experts. Meanwhile, the public, left without clear recommendations, was left to react on an emotional level about the pandemic as death and other severe clinical complications occurred because of the lack of common sense. Nobody shows up well. Regulators, usually wiser, have not been able to be a voice of reason heard by the public. While the FDA has faced a difficult situation dealing with direct and indirect pressure from the US president, the EMA has been relatively absent from this discussion.

Assignment of blame is not necessary here, but we do need to learn from the situation for the future. Obviously, there will not be a united voice worldwide as individual countries try to gain advantage over others. WHO’s credibility has been damaged, and it will not have a strong voice for several years while they undergo some inevitable reform. Finally, the EU may yet consider this failure to manage the COVID-19 crisis and learn from it for the future. But the only certain thing is that a year from now priorities will shift.

## Data Availability

All data in the manuscript were available.

## Declarations

### Funding

the authors received no financial support for the research, authorship and/or publication of this article.

### Conflict of interest

The authors declare that they have no conflicts of interest.

### Ethic approval

not required as this study used China National Knowledge Infrastructure, ClinicalTrials.gov, the EU Clinical Trials Register, and the Chinese Clinical Trial Register databases

Consent to participate not applicable to this study

Consent for publication: not applicable to this study

Availability of data and material (data transparency): not applicable to this study

### Code availability

not applicable to this study

### Authors’ contributions

Mondher Toumi, Yitong WANG and Shuyao LIANG contributed to the study conception, design, data curation and original draft preparation. Tingting QIU and Ru HAN contributed to the study conception, design, data curation and review of the article. Monique DABBOUS contributed to editing and review of the article. All authors read and approved the final manuscript.

## Acknowledgments

The authors thank Emilie Clay, of Creativ-Ceutical, who provided an insightful review and comments on the manuscript. Medical editorial assistance was provided by Steven Melvin of ApotheCom (Yardley, PA, USA).

## Notes

### Competing Interest Statement

The authors have declared no competing interest.

### Author Declarations

The IRB/oversight body is an exemption for the research.

